# Distinct metabolic signatures of Alzheimer’s and Parkinson’s disease revealed through genetic overlap with metabolic markers

**DOI:** 10.1101/2025.07.31.25332114

**Authors:** Sara E. Stinson, Alexey A. Shadrin, Zilur Rahman, Linn Rødevand, Iris J. Broce, Karin Persson, Geir Selbæk, Hreinn Stefansson, Jan Haavik, Nadine Parker, Elise Koch, Oleksandr Frei, Kevin S. O’Connell, Olav B. Smeland, Srdjan Djurovic, Anders M. Dale, Dennis van der Meer, Ole A. Andreassen

## Abstract

Metabolic dysfunction is increasingly implicated in neurodegenerative diseases, yet the genetic architecture linking metabolic markers with Alzheimer’s disease (AD) and Parkinson’s disease (PD) remains unclear. We systematically analysed phenotypic and genetic relationships between 249 circulating metabolites with AD and PD, comparing patterns to body mass index (BMI), type 2 diabetes (T2D), coronary artery disease (CAD) and stroke. Using linkage disequilibrium score regression and bivariate Gaussian mixture modeling, we identified distinct genetic overlap. AD correlated positively with cardiometabolic traits (BMI, r_s_=0.11; T2D, r_s_=0.23; CAD, r_s_=0.22; stroke, r_s_=0.18), whereas PD showed opposing patterns (AD–PD r_s_=−0.36). Mendelian randomization identified bi-directional causal effects of lipid measures on AD and divergent effects of glutamine on AD and PD. Conjunctional FDR analyses mapped 1,377 shared genes, implicating lipid metabolism in AD and synaptic processes in PD. These findings disentangle disease-specific pathways and inform therapeutic strategies targeting metabolic health.

## INTRODUCTION

The prevalence of neurodegenerative diseases (NDDs) is expected to rise sharply as global life expectancy increases, contributing to profound societal and economic burdens^1^. Alzheimer’s disease (AD), followed by Parkinson’s disease (PD) are the two most common forms of NDDs^2^. Unlike other leading causes of death, such as coronary artery disease (CAD) which have plateaued or even declined in incidence, the prevalence of NDDs continues to rise at an alarming rate^3^.

Dysregulation of metabolic pathways^4^ and immune-related factors^5^ have emerged as key contributors to neurodegeneration. Cardiometabolic diseases (CMDs), including obesity, type 2 diabetes (T2D), CAD, and stroke are established risk factors for accelerated cognitive decline, with lifestyle factors such as physical inactivity, smoking, and excessive use of alcohol, compounding these effects^6^. These associations have been extensively studied in AD^7^, with T2D and elevated blood glucose levels increasing the risk of AD^8,9^. Overweight and obesity in midlife are similarly associated with heightened AD risk^10–12^. Parallel findings have also emerged for PD, where T2D is associated with higher risk of PD and faster progression of symptoms^13–15^.

Beyond epidemiological associations, genetic evidence supporting biological links between CMDs and NDDs remains mixed, with some studies reporting a lack of significant genetic correlations^16^. Yet several studies report shared genetic architecture between T2D and AD, with several genome-wide association studies (GWASs) identifying common single-nucleotide polymorphisms (SNPs) across conditions^17–19^. Many of these shared loci converge on pathways related to lipid metabolism, insulin-resistance, immune response, cell signalling, and neuronal plasticity^18,20^. The *APOE* (encoding apolipoprotein E) ε4 allele, the strongest genetic risk factor for late-onset AD, plays a central role in lipid metabolism and cholesterol transport, contributing to both cerebrovascular dysfunction and amyloid pathology^21^. The *APOE* locus has also been implicated in PD, influencing age of onset and the development of PD-related dementia^22^. However, despite some overlapping risk loci between AD and PD, the overall pairwise genetic correlation is low (r_g_=0.14, p=0.07)^16^. Additional PD-associated genes, such as *LRRK2*, *GBA,* and *PINK1* indicate involvement of mitochondrial dysfunction, lysosomal activity, and lipid metabolism^23^.

Despite significant advances in understanding the mechanisms behind NDDs, effective disease-modifying treatments remain scarce^24^. Current therapies, such as levodopa (L-DOPA) for PD, a dopamine precursor, provide only symptomatic relief^25^. Likewise, treatments for AD, including acetylcholinesterase inhibitors and NMDA receptor antagonists, have been available for decades but primarily manage symptoms. Although many compounds targeting amyloid-β plaques have failed in late-stage trials, several recent drugs have been approved that modestly slow cognitive decline^26^. In contrast, therapies developed for CMDs, including those for T2D, show promise due to their pleiotropic effects on multiple metabolic and inflammatory pathways^27^. Notably, clinical trials of GLP-1 receptor agonists (GLP1-RAs), which induce glucose-stimulated insulin secretion, but also prompt significant weight loss, seem to exert neuroprotective effects in AD, with some evidence in PD^28,29^. It is also estimated that 40% of dementia cases may be preventable through modifiable risk factors, such as physical inactivity, smoking, alcohol intake, obesity, and T2D^30^, underscoring the potential of interventions targeting improved metabolic health.

While mounting evidence implicates metabolic dysfunction in NDDs^7^, critical questions remain, regarding shared and distinct biological pathways across AD and PD, and their intersection with CMDs. Advances in high-throughput metabolomics, particularly nuclear magnetic resonance (NMR) spectroscopy^31^, now enable comprehensive profiling of circulating lipids, lipoproteins, fatty acids, amino acids, glucose, and inflammatory metabolites in large scale biobanks. Metabolomics shows strong promise for AD risk prediction, with age, sex, and metabolic profiles outperforming established clinical predictors; however, this advantage was not observed for PD^32^. Moreover, we have recently conducted the largest GWAS of circulating metabolic markers in 328,006 individuals, identifying up to 15,585 loci (465 unique genomic regions; 166 of which were novel)^33^, offering unique opportunities to elucidate the metabolic underpinnings of neurodegeneration.

Here, we systematically investigate the genetic overlap between NDDs and circulating metabolic markers, and compare these association profiles with CMDs, to reveal shared genetic architecture and infer causality. We apply linkage disequilibrium score regression (LDSC) to quantify global genetic correlations, bivariate Gaussian mixture modelling (MiXeR) to assess polygenic overlap, and perform Mendelian randomization (MR) to evaluate causal relationships. Furthermore, we employ conjunctional false discovery rate (conjFDR) analysis to identify shared genetic loci between traits, followed by gene set enrichment analyses to uncover functional pathways, and tissue-specific gene expression for biological relevance of implicated genes. An overview of the study design is provided in **Fig 1**. We hypothesize that AD and PD will exhibit distinct metabolic signatures, reflecting disease-specific etiologies. This systematic genetic approach allows us to deconstruct the metabolic architecture of neurodegeneration, revealing disease-specific pathways that are essential for developing targeted therapeutic strategies.

**Fig 1.**
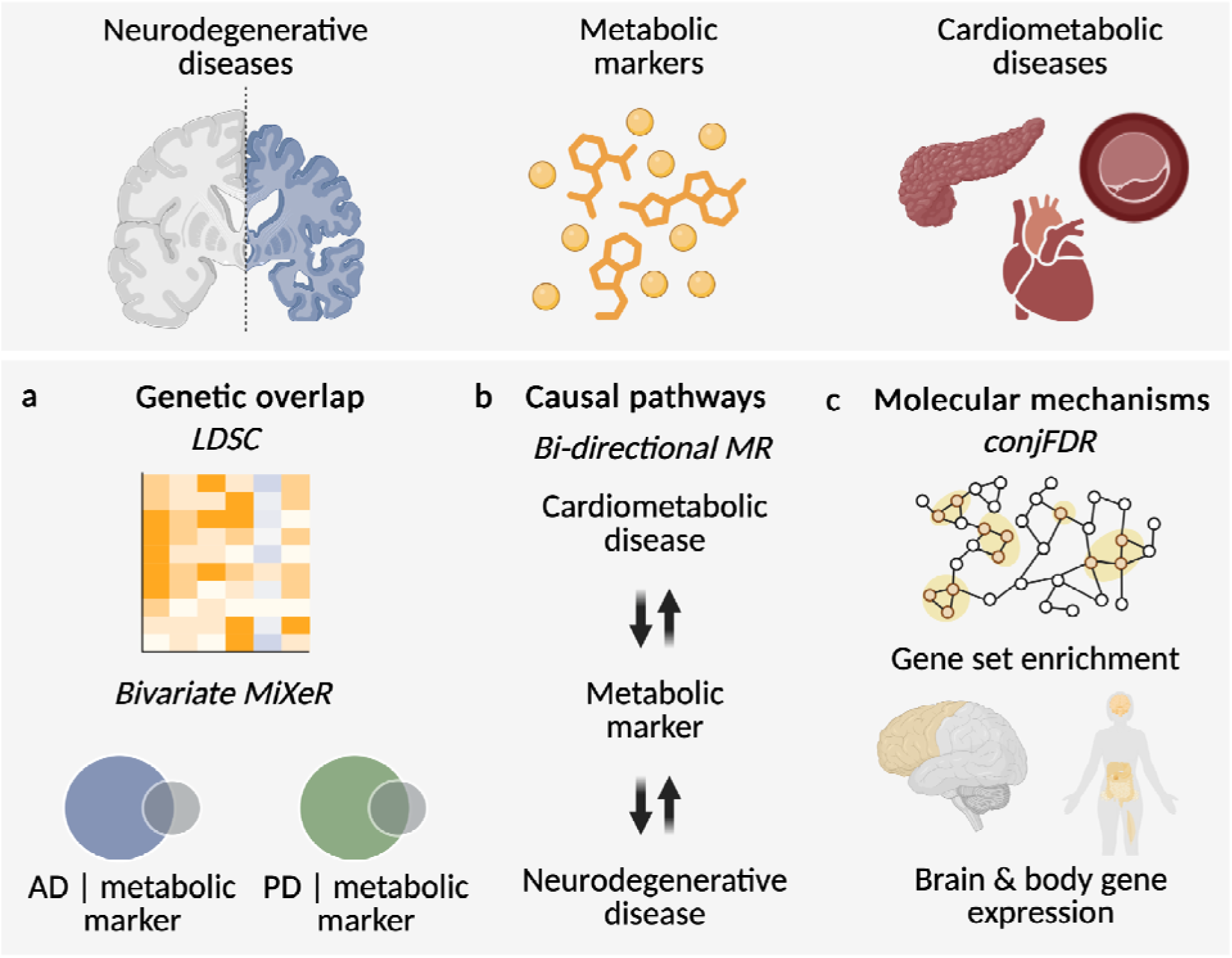
Overview of study design. This study investigates the genetic overlap between metabolic markers, neurodegenerative diseases (NDDs), and cardiometabolic diseases (CMDs). **a,** Global genetic overlap was measured using linkage disequilibrium score regression (LDSC) and bivariate Gaussian mixture modelling (MiXeR). **b,** Potential bidirectional causal relationships between metabolic markers, NDDs and CMDs were evaluated using Mendelian randomization (MR). **c,** Shared genetic loci between metabolic markers and NDDs were identified through conjunctional false discovery rate (conjFDR) analysis, followed by gene set enrichment and tissue-specific (body and brain) gene expression analyses to explore functional pathways and biological relevance (Figure generated with <u>Biorender.com</u>).

## RESULTS

### Phenotypic overlap

We first explored phenotypic associations between metabolic markers and traits of interest, in 207,841 unrelated UK Biobank participants of European ancestry with plasma concentrations of 249 metabolic markers from the Nightingale NMR panel. This panel includes 228 lipids, lipoproteins, or fatty acids, and 21 non-lipid traits such as amino acids, ketone bodies, and glycolysis- and inflammation-related markers (see **Suppl Table 1** for complete list). Disease diagnoses were derived from ICD-10 codes, for AD (n=1,746 cases), PD (n=1,910), T2D (n=17,477), CAD (n=25,705), and stroke (n=8,681). BMI was analyzed as a continuous trait (mean 27.4, standard deviation [SD] = 4.8). Associations were estimated via linear or logistic regression, adjusted for age and sex.

We observed extensive and highly significant associations between metabolic markers and NDDs (**Suppl Fig 1a**). Metabolites most strongly associated with higher AD risk, included higher levels of glucose, S-LDL-PL%, and M-LDL-PL%, and lower levels of valine, total branched-chain amino acids (BCAA), and omega-3 fatty acids. For PD, key metabolites included higher levels of S-LDL-PL%, IDL-PL%, and glycine, and lower levels of S-HDL-FC, omega-6, and S-HDL-C (all *p*<0.05 FDR). The overall phenotypic association patterns between metabolic markers and AD vs. PD were moderately correlated (r_s_=0.58; **Suppl Fig 1b**). Notably, an inverse pattern of associations was observed for BMI with PD (r_s_=-0.32) and AD (r_s_=-0.18). Comparative patterns of phenotypic associations of 249 metabolic markers with AD and PD, in comparison to CMDs, is visualized by the heatmap in **Suppl Fig 1c**.

### Genetic overlap

To assess genetic overlap, we used GWAS summary statistics generated from the Nightingale panel of 249 metabolic markers^33^. GWAS summary statistics were also obtained for AD^34,35^ and PD^36,37^, major CMDs including T2D^38^, CAD^39^ and stroke^40^; and body mass index (BMI)^41^ as a key cardiometabolic risk factor (**Suppl Table 2**).

Using LDSC^42^, we estimated genetic correlations (r_g_) between each metabolic marker and disease traits. This revealed widespread genetic associations between metabolic markers and NDDs, although the extent and direction of associations differed between AD and PD. A summary of genetic correlations is presented in **Fig 2a**, with volcano plots highlighting the strongest associations with metabolic markers and NDDs. Overall, we identified nominally significant genetic correlations (*p*<0.05) for 17 out of 249 markers (6.8%) with AD and 4 markers (1.6%) with PD; though none passed multiple testing correction, false discovery rate (FDR)<0.05. This revealed a key divergence, as the overall genetic correlation patterns for metabolic markers between AD and PD were negatively correlated (Spearman’s r_s_=-0.36), suggesting opposing metabolic-genetic influences. This genetic opposition stands in sharp contrast to their positive phenotypic association pattern (r_s_=0.58, **Suppl Fig 1b**). This is distinct from the known lack of significant genetic correlation between the two NDDs (AD-PD, r_g_=0.13; *p*=0.15), see **Fig 2b**. When comparing the patterns of metabolic marker associations of AD and PD to CMDs, a clear divergence emerged, visualized by the heatmap in **Fig 2c**. Specifically, AD displayed more similar patterns of genetic correlations across the markers to BMI, T2D, CAD, and stroke, while PD showed strikingly opposite directions of correlations. These results suggest that some of the metabolic pathways involved in AD etiology may differ from those in PD, and that the genetics of metabolic processes may help distinguish between NDDs. Full results are listed in **Suppl Table 3**.

**Fig 2.**
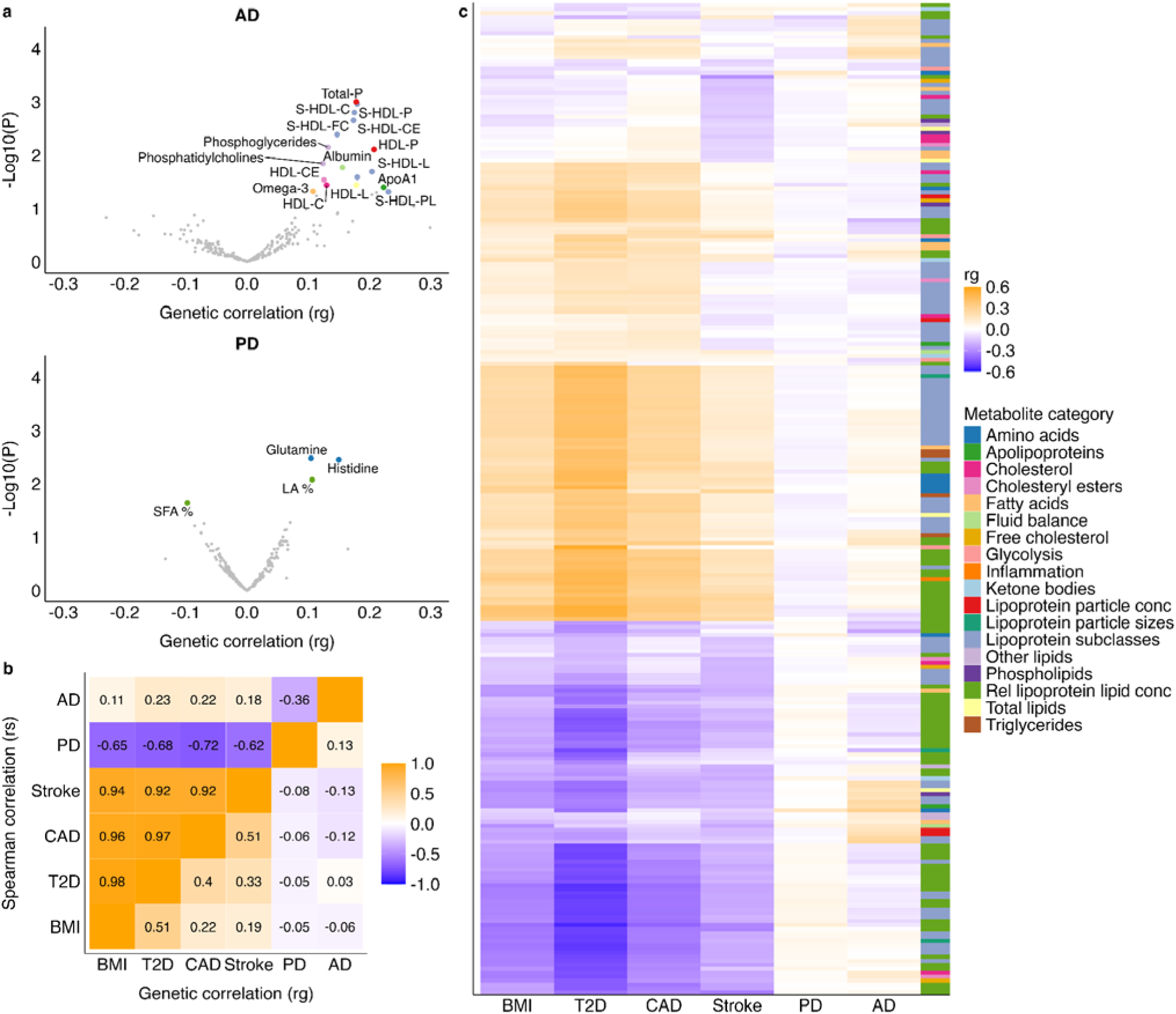
Genetic correlations with metabolic markers, AD, PD and cardiometabolic disease/traits (LDSC). **a,** Volcano plots summarizing the genetic correlations on the x-axis and the -log10(p-values) on the y-axis for AD and PD. Metabolic markers with nominally significant correlations (p ≤ 0.05) are colored according to their metabolic category and annotated with their names, while non-significant markers (p>0.05) are shown in gray. **b,** Correlation matrix comparing the genetic correlation estimates across traits. The bottom right triangle displays pairwise genetic correlations (rg), while the upper left triangle shows Spearman correlation coefficients of LDSC estimates across all 249 metabolic markers. **c,** Heatmap of genetic correlations between 249 metabolic markers (y-axis) and cardiometabolic disease traits (BMI, T2D, CAD, and stroke) as well as AD and PD (x-axis). Positive correlations are shown in orange, while negative correlations are in blue (see legend). The metabolic markers (y-axis) are hierarchically clustered, with categories indicated in the adjacent color bar. Overall, the plots reveal a divergent pattern, where the metabolic-genetic profile of AD shows similarity to cardiometabolic traits, while the profile for PD is largely opposing. AD=Alzheimer’s disease; BMI=body mass index, CAD=coronary artery disease, PD=Parkinson’s disease, T2D=type 2 diabetes.

The polygenicity of metabolic markers, defined as the number of SNPs influencing each trait using univariate MiXeR^43^, ranged from 17 (SD=3) to 1,529 (SD=52), with a mean of 261 (SD=26) SNPs per metabolite, as previously reported^33^. For the NDDs, 345 (SD=53) SNPs influenced AD and 875 (SD=116) SNPs for PD, reflecting the higher polygenicity of PD^16^.

To extend beyond genetic correlations^42^, we applied bivariate MiXeR^44^ to quantify the fraction of trait-influencing variants common to metabolic markers and NDDs, irrespective of effect direction (**Fig 3a**). Model fit improved over the reference model for 236 of 249 metabolites (94.8%) in AD and 219 of 249 metabolites (88.0%) in PD.

**Fig 3.**
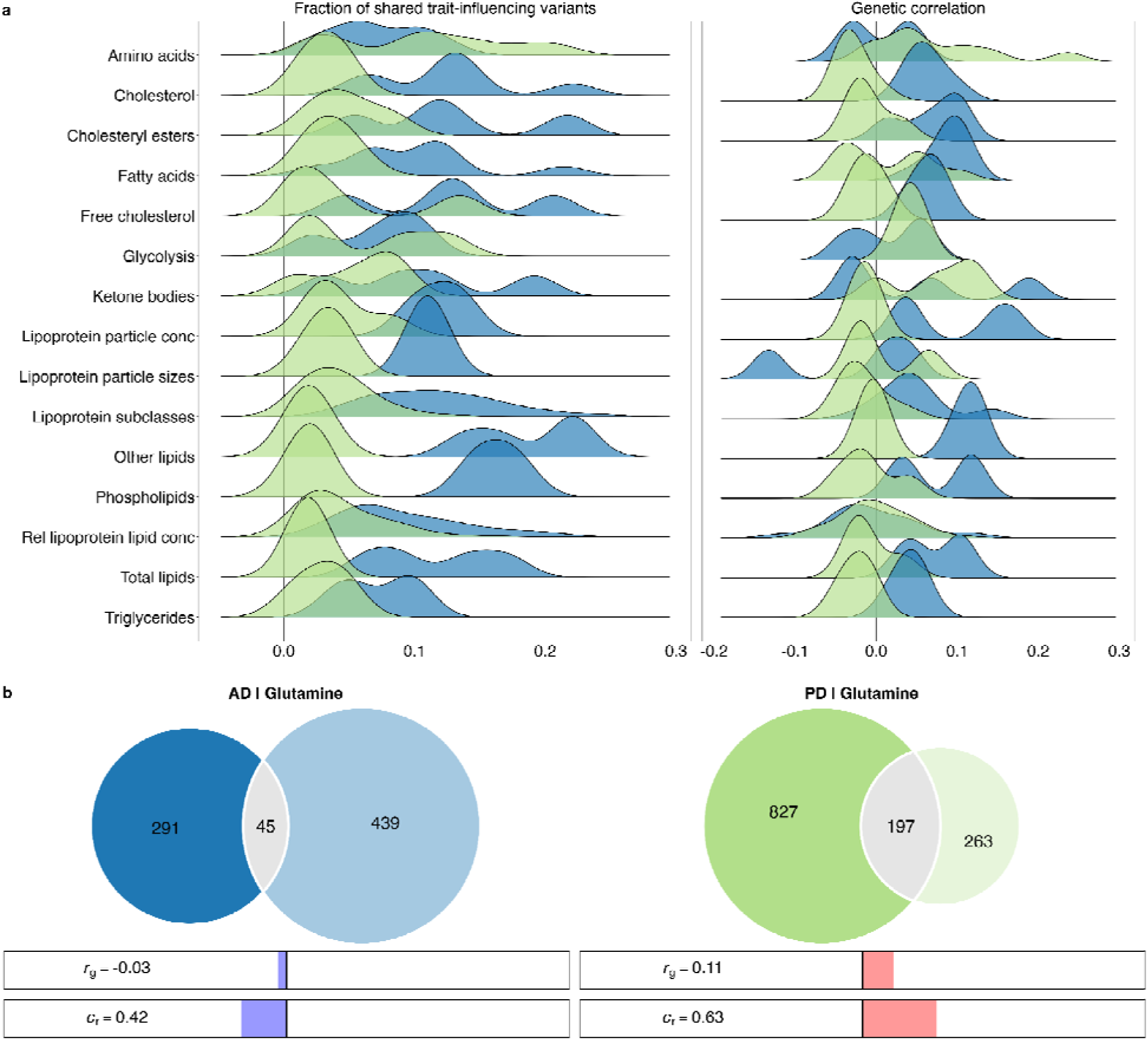
Genetic overlap between metabolic markers and NDDs. **a,** Density plots show the distributions of the fraction of shared trait-influencing variants (left) and genetic correlations (right) from bivariate MiXeR, across metabolite categories (>1 metabolite per category). **b,** Venn diagrams illustrating the estimated number of unique and shared variants influencing glutamine, AD and PD. Below the Venn diagrams, the bar displays the genetic correlation (r_g_) and concordance rate (C_r_; wherein 0.50 indicates an equal number of variants with opposing and same directions of effects on the pair of traits). AD=Alzheimer’s disease (blue); PD=Parkinson’s disease (green).

AD exhibited a significantly greater proportion of shared variants with metabolic markers than PD (median: 10.5% vs. 3.6%; range: 0.6–24.3% vs. 0.3–21.1%; Wilcoxon p=3.5×10^-30^). These overlaps occurred alongside modest genetic correlations (AD: median r_g_=0.04, range: −0.13–0.19; PD: median r_g_=−0.02, range: −0.09–0.24), driven by intermediate concordance rates (AD: median c_r_ = 0.56, range: 0.12–0.91; PD: median c_r_ = 0.44, range: 0.06–0.94), suggesting that although shared variants are common, their effect directions are often mixed.

Among the metabolic categories, other lipids, phospholipids, and free cholesterol had the highest proportion of shared variants with AD (median: 19.3%, 16.2% and 12.9%, respectively). Marked examples for AD include S-HDL-FC (24.3%), S-HDL-P (23.9%) and XL-HDL-CE (23.5%; **Suppl Fig 2a**). In contrast, amino acids, fluid balance and ketone bodies exhibited the highest overlap of shared variants with PD (median: 10.9%; 6.6% and 6.5%, respectively). Notable amino acids for PD include, leucine (21.1%), valine (18.6%), and glutamine (15.3%; **Suppl Fig 2e**).

Glutamine serves as a remarkable example of a metabolite with an opposing pattern of genetic overlap (AD: r_g_=-0.03, PD: r_g_=0.11; **Fig 3b**). The proportion of shared variants was greater for PD (15.3%) than for AD (5.8%). Although this difference could be in part due to the higher polygenicity of PD, with a substantially larger number of causal variants compared to AD. Comprehensive overlap estimates for all metabolite– disease pairs are presented in **Suppl Table 4**, with additional distinct examples presented in **Suppl Fig 2.**

### Causal relationships

We performed bidirectional MR to identify significant causal relationships between metabolic markers and NDDs. **Fig 4** presents the Inverse Variance Weighted (IVW) MR coefficients that remained significant after correction for multiple comparisons, and were also significant through Weighted Median and MR Egger approaches, confirming the robustness of the findings (see **Methods**).

**Fig 4.**
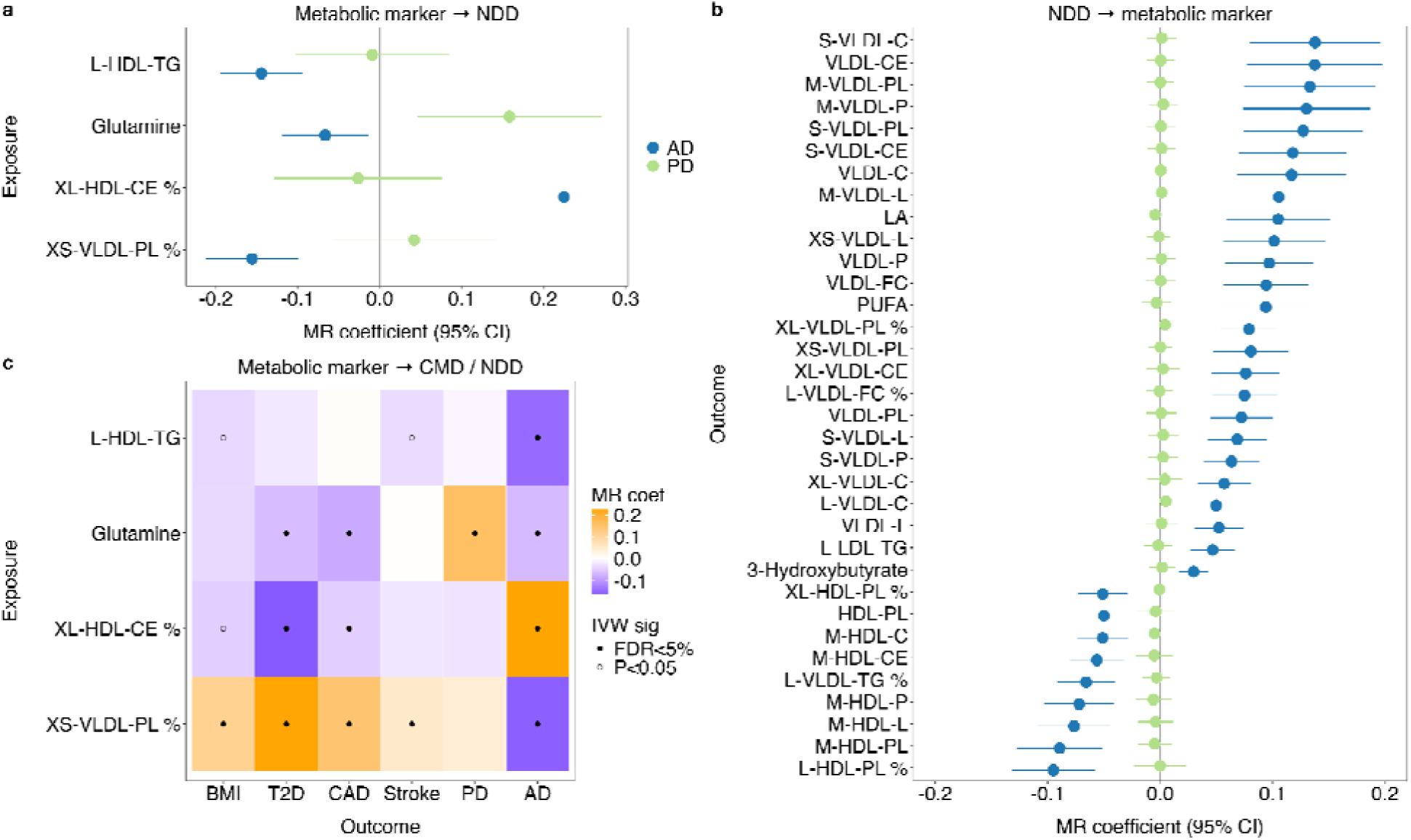
Bidirectional causal relationships between metabolic markers, NDDs, and CMDs. **a,** Forest plot of the causal effects of significant metabolic markers on NDDs (AD, PD), with IVW MR coefficients and their 95% CIs on the x-axis. **b,** Forest plot of the causal effects of NDDs on top metabolic markers (FDR<1.0×10⁻□). For both panel a and b, the metabolic markers are shown on the y-axis and the two NDDs are color-coded in the legend. **c,** Heatmap of the significant metabolic markers on NDDs, and their direction of effects on BMI and CMDs. AD=Alzheimer’s disease; BMI=body mass index; CAD=coronary artery disease; CIs=confidence intervals; CMD=cardiometabolic disease; IVW=Inverse Weighted Median; MR=Mendelian randomization; NDD=neurodegenerative disease; PD=Parkinson’s disease; T2D=type 2 diabetes.

As shown in **Fig 4a**, four metabolic markers (L-HDL-TG, glutamine, XS-VLDL-PL%, and XL-HDL-CE%) had causal effects on AD, whereas two markers demonstrated suggestive causal effects on PD (glutamine and leucine; FDR<0.05 for IVW and MR Egger only, respectively). See **Suppl Fig 3** for comparison of significant metabolic markers on NDDs across MR methods. **Fig 4b** shows the reverse direction, further highlighting the distinction between the two NDDs, where AD exhibited widespread robust causal effects on 179 metabolic markers, of which the majority were lipids, compared to PD with no significant causal associations with metabolic markers. AD had widespread causal effects across metabolic marker categories, notably for cholesterol subtypes and apolipoproteins (See **Suppl Fig 4**). When we compared the metabolic markers casual for AD, interestingly we observed similar directions of causal effects of glutamine on AD, as with T2D and CAD on one side and PD on the other side, as represented in **Fig 4c**. Bidirectional MR results between metabolic markers and diseases are presented in **Suppl Tables 5-6**.

We subsequently expanded the MR analyses to investigate the causality between BMI and CMDs on NDDs. We found no robust evidence of direct causal effects of these cardiometabolic traits on the NDDs, despite their well-known phenotypic relationships. BMI and CMDs had bidirectional causal relationships with the metabolic markers. AD had more bidirectional causal relationships with the markers than PD (**Fig 4a-b**). This may suggest that the metabolic markers are intermediates between cardiometabolic health and neurodegeneration. MR results of cross-disease pairs are provided in **Suppl Tables 7**.

### Molecular mechanisms

To investigate biological pathways linking NDDs and metabolic health, we performed conjFDR analyses^45^. This approach identified 1,282 genetic variants for AD and 145 for PD jointly associated with 249 metabolic marker-trait pairs (**Suppl Tables 8-9**). Among the metabolic markers with suggestive causal effects on NDDs, conjFDR highlighted 153 loci shared by AD and XL-HDL-CE % (**Suppl Fig 5a**), 14 loci for AD and glutamine (**Suppl Fig 5b**), 140 loci for AD and L-HDL-TG (**Suppl Fig 4c**), 167 loci associated with AD and XS-VLDL-PL % (**Suppl Fig 5d**), 15 loci for PD and glutamine (**Suppl Fig 5e**), and 1 locus for PD and leucine (**Suppl Fig 5f**). Notably, the AD-glutamine analysis identified rs2840676 in *LEPR*, previously linked to BMI, schizophrenia, and immune traits^46,47^. For PD-glutamine, notable significant loci included rs11223625 (*IGSF9B*), associated with lower waist circumference and body fat percentage^48^, and rs11614702 (*FBRSL1*), linked to glucose metabolism, T2D, and statin response^49^, and rs2274693 (*PTCH1*), associated with cortical surface area^50^.

The shared lead variants were mapped to genes using OpenTargets^51^, and these genes were tested for enrichment in Gene Ontology (GO) biological processes. **Fig 5a** shows the proportion of shared and distinct aggregated genes identified through the conjunction of NDDs with metabolic markers. **Fig 5b** lists the top significant GO terms for each NDD among the 249 marker pairings. For the AD-marker pairs, lipid metabolism and transport pathways were prominently enriched; while pathways related to synaptic function were more common for PD. The two NDDs exhibited only a small number of shared variants (n=29), leading to mostly distinct biological processes that mediate their relationship with metabolic markers. **Suppl Tables 10-11** provide a comprehensive overview of all significant GO terms and their enrichment, per disease.

**Fig 5.**
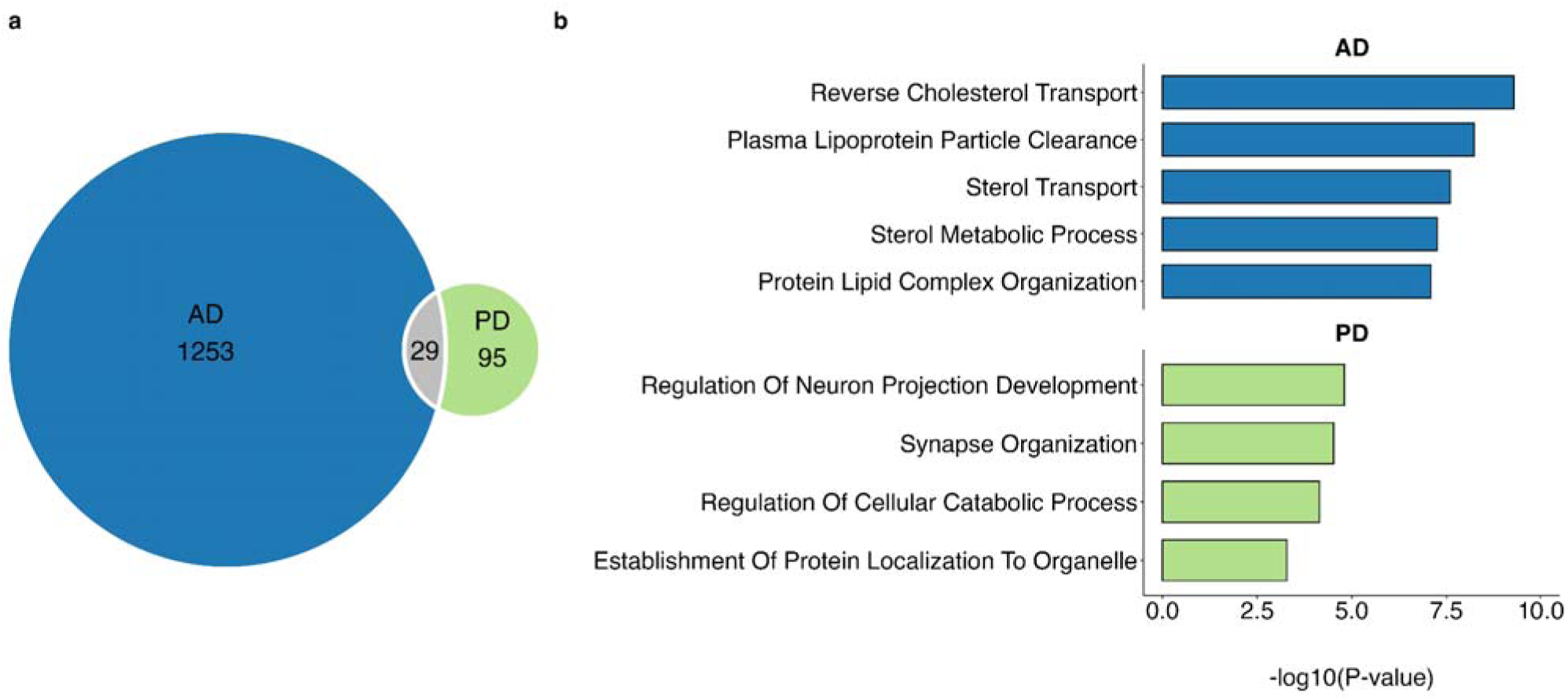
Coupling of genes between metabolic markers with AD and PD to gene ontology terms. **a,** Venn diagram depicting the number of unique and shared genes found through conjFDR analyses, aggregated across all markers, for AD and PD. **b**, Plots showing the top most significantly enriched gene ontology terms (y-axis), based on the mapped genes in AD and PD (panels)), with the observed p-values (x-axis). AD=Alzheimer’s disease; cFDR=conjunctional false discovery rate; PD=Parkinson’s disease.

To assess the relevance of our findings for brain and body health, we linked the two aggregated sets of mapped genes identified through the conjFDR approach to tissue gene expression data. For both AD and PD, there was enrichment across multiple brain regions and body tissues, with differing profiles. **Fig 6a** illustrates regional variation in gene expression across cortical and subcortical brain regions. For AD, suggestive enrichment was observed exclusively in the superior frontal cortex region (*p*=0.0480, FDR>0.05). For PD, suggestive enrichment was found in the caudate (*p*=0.0198), cuneus (*p*=0.0205), caudal anterior cingulate (*p*=0.0259), pericalcarine (*p*=0.0358), and superior frontal regions (*p*=0.0359) (all FDR>0.05). **Fig 6b** shows the enrichment across various body tissues as per GTEx v8. These results indicate that the mapped genes are not solely expressed in the brain (*FDR*=1.69×10^-23^) for AD, but are also enriched across the whole-body, most significantly in tissues such as the pancreas (*FDR*=3.66×10^-27^), liver (*FDR*=3.66×10^-27^) and heart (*FDR*=3.53×10^-26^). For PD, the non-central nervous system (CNS) tissue enrichment was less prominent, with the heart (*FDR*=1.16×10^-6^), liver (*FDR*=1.16×10^-4^), skeletal muscle (*FDR*=0.002), and the pancreas (*FDR*=0.003) most significantly involved.

**Fig 6.**
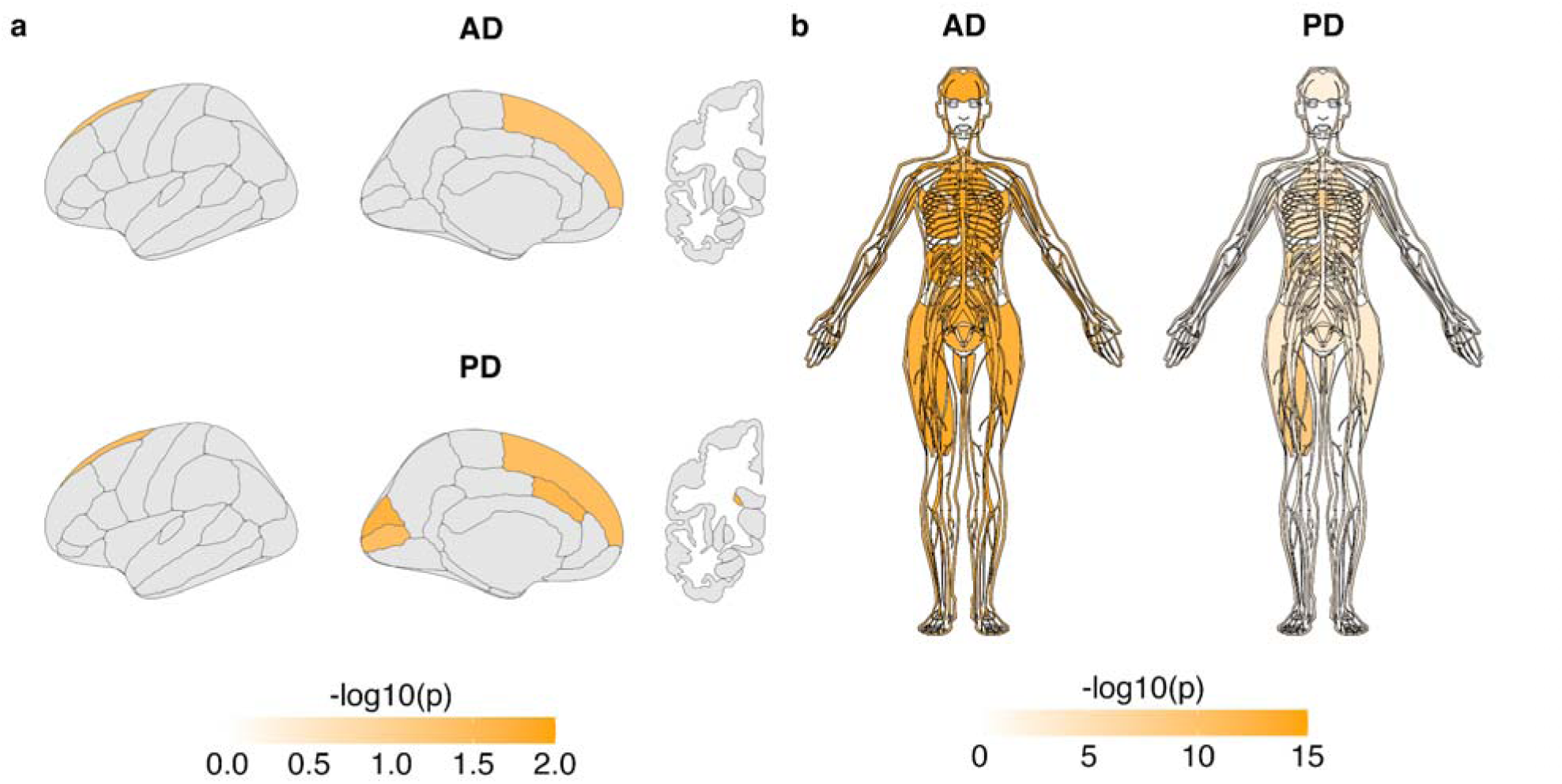
Expression patterns of genes shared between metabolic markers and AD and PD across body and brain tissues. **a,** Brain maps displaying the significance (p<0.05) of enrichment tests across cortical regions (Desikan-Killiany atlas) and subcortical regions (Freesurfer aseg), based on expression data from the Allen Brain Atlas. **b,** Antograms visualising the results of the enrichment tests for each of 30 general tissue types from the GTEx v8 database. Colour coding indicates significance as -log10(p-value) of enrichment tests assessing the overlap of shared genes in AD and PD. AD=Alzheimer’s disease; PD=Parkinson’s disease.

## DISCUSSION

Our findings demonstrate that while metabolic pathways are broadly implicated in NDDs, their genetic signatures are highly distinct for AD and PD, with divergent links to CMDs and BMI. Most notably, we observed distinct differences in the shared genetic loci and architecture of metabolic markers with AD vs. PD, despite their similar phenotypic associations with these markers. MR analyses revealed robust bi-directional causal effects between metabolic markers and AD, but only suggestive uni-directional effects of metabolic markers on PD. Functional annotation of shared genetic variants highlighted lipid metabolism and transport for AD, and neuronal activity for PD, also supported by the whole-body gene expression results.

These divergent metabolic signatures have important clinical implications. The contrasting profiles of AD and PD suggest that genetically informed metabolic biomarkers could aid early differential diagnosis of neurodegenerative diseases. Furthermore, while metabolic interventions may hold promise for AD, they may offer limited benefit, or even potentially detrimental in PD, underscoring the need for precision approaches. Developing disease-specific metabolic biomarker panels and targeted therapies represents a critical step away from one-size-fits-all strategies toward tailored prevention of neurodegeneration.

While AD and PD exhibit a relatively low degree of genetic overlap^34^, our results show that their genetic relationships with metabolic markers are distinctly divergent, contrasting their phenotypic similarities. Specifically, the genetic architecture of AD appears more closely aligned with CMDs, while PD exhibits an opposing pattern. While prior studies have found inconsistent direct genetic associations between BMI and T2D on AD^52^, epidemiological evidence suggests that higher midlife BMI increases dementia risk, whereas higher late-life BMI may confer protection^11^. Our genetic correlation patterns between metabolic markers and AD mirrored those of BMI, yet were opposite in direction to their phenotypic associations. These findings support the hypothesis that age-dependent metabolic shifts occur during mid-life, followed by compensatory mechanisms in later life, which may alter these relationships over the lifespan^53^.

Metabolic dysregulation has been previously implicated in NDDs, including disruptions in glucose metabolism, lipid homeostasis, amino acid signaling, and neuroinflammatory processes^7,23,54,55^. Understanding causal pathways linking metabolism and neurodegeneration has potential to identify novel therapeutic strategies. Previous MR studies have implicated specific metabolites, such as glutamine in AD and leucine in PD^56^. Our findings provide evidence for robust causal relationships between several circulating metabolic markers and AD, particularly those related to lipid metabolism, and suggestive evidence for PD. Notably, bidirectional effects were prominent for AD, indicating a complex interplay where metabolic dysregulation may not only contribute to AD onset but also be influenced by neurodegenerative disease processes, which is important for the development of interventions.

We identified a causal association between lower circulating glutamine levels and higher AD risk, mirroring the pattern observed for cardiometabolic diseases (including CAD^57^), and contrasting with the opposite effect in PD. Glutamine serves as a precursor for glutamate, the primary excitatory neurotransmitter essential for synaptic plasticity and cognitive function through NMDA receptor activation. Disruptions in glutamatergic signalling have long been implicated in AD pathology, with NMDA receptor antagonists currently used to treat cognitive symptoms in AD^58^. Yet, GWASs of BMI have also implicated glutamatergic signaling and NMDA receptor-related neuroplasticity in body weight regulation^41^. Moreover, recent preclinical studies suggest that combining NMDA receptor antagonists with GLP1-RAs, agents already known for their metabolic benefits, could hold added therapeutic potential for obesity treatment^59^. GLP1-RAs exhibit neuroprotective properties^60^, highlighting possible avenues for targeting both metabolic and neurodegenerative pathways simultaneously, reinforcing the metabolic contribution to AD pathogenesis.

In addition to glutamine, lipids emerged as a key area of bidirectional interaction with AD. The MR analyses of several HDL- and VLDL-related metabolites showed robust causal effects on AD risk, reinforcing the central role of dysregulated lipid processing in neurodegeneration. These findings align with existing knowledge around *APOE*, a major genetic risk factor for late-onset AD, which is intimately involved in lipid transport and metabolism^61^. The observation that AD exerted causal effects on lipid traits, but not for PD, may support distinct metabolic signatures underpinning these two NDDs.

By applying a conjFDR approach, we identified hundreds to thousands of genetic variants jointly associated with NDDs and circulating metabolic markers, providing new insights into the molecular pathways linking metabolic health and neurodegeneration. Our pathway analyses revealed distinct biological signatures for AD and PD. In AD, the pathways predominantly involved lipid metabolism and transport, particularly cholesterol and sterol-related processes, which are critical for amyloid-β clearance, *APOE* function, and maintaining neuronal membrane integrity. These findings align with longstanding evidence implicating dysregulated lipid metabolism in AD pathogenesis^61^. For PD, the enriched pathways were related to neuronal development and maintenance, including synaptic organization, neuronal projection, and protein localization, processes which are essential for the survival and function of dopaminergic neurons and the regulation of protein aggregation^62^. Together, these results underscore the distinct molecular mechanisms, in which metabolic dysregulation may contribute to differences in NDDs.

Narrowing our conjFDR analysis to metabolites with potential causal effects revealed specific genetic loci that suggest novel mechanistic links between metabolism and neurodegeneration. For AD and glutamine, the lead variant rs2840676 mapped to *LEPR*, a key regulator of leptin signalling, insulin sensitivity, and energy homeostasis, previously associated with BMI, inflammation, and schizophrenia^46,47^. These findings underscore the convergence of metabolic and inflammatory pathways in AD vulnerability, consistent with evidence linking impaired leptin signalling to amyloid pathology and neurodegeneration^63^. For PD and glutamine, the associated loci included *IGSF9B*, implicated in synaptic adhesion and lower BMI, and *FBRSL1*, a gene linked to glucose metabolism and T2D. Together, these findings position glutamine metabolism as a common yet directionally opposing axis that connects systemic metabolic states to neuronal integrity via disease-specific mechanisms.

We mapped the genes implicated through conjFDR to tissue-specific expression profiles across the brain and peripheral organs. In the brain, genes associated with AD showed suggestive enrichment in the superior frontal cortex, a region critical for executive functions and among the earliest affected in AD-related cognitive decline^64^. For PD, genes were enriched in several regions including the caudate, cuneus, caudal anterior cingulate, pericalcarine, and superior frontal areas, consistent with known PD pathology affecting motor control, sensory processing, and executive function^22^. Notably, our findings of enrichment in cortical regions such as the superior frontal cortex (AD and PD) and cuneus (PD), may align with the reported expression of incretin receptors (GLP-1R and GIPR) throughout the cerebral cortex^27^. This partial overlap suggests that some of the brain regions responsive to incretin-based therapies may partly mediate metabolic influences on neurodegeneration and should be further explored.

Importantly, the enriched gene expression was not restricted to the brain. In AD, we observed strong enrichment in peripheral metabolic organs, including the liver, pancreas, and heart, underscoring the systemic nature of AD. This pattern reinforces the hypothesis that chronic systemic metabolic dysfunction may impose a long-term “metabolic overload” on the brain^65^. Such overload could disrupt insulin and lipid signaling, impair amyloid-β clearance, and compromise neuronal energy homeostasis, ultimately lowering the threshold for amyloid plaque formation, tau hyperphosphorylation, and neuronal death^7^. In PD enrichment was also evident in skeletal muscle, heart, liver, and pancreas, though to a lesser extent than in AD. Peripheral involvement in skeletal muscle may reflect pathways linked to motor dysfunction, autonomic regulation, and muscle rigidity, hallmarks of PD^23^. The accumulation of α-synuclein within Lewy bodies creates a membrane-rich environment characterized by abnormal vesicular structures, dysfunctional mitochondria, and elevated lipid content^66^, aligning with the view of PD as a multisystem disorder with early pathology outside the CNS^67,68^.

A major strength of our study lies in its large-scale, integrative approach, leveraging comprehensive genetic, metabolic, and functional annotation analyses to unravel shared and distinct molecular mechanisms underlying AD and PD. Additionally, we leveraged the latest and most well-powered GWAS for NMR metabolomics in over 300 thousand individuals^33^. However, an inherent limitation of this study is the unequal statistical power of the GWASs for the two NDDs, coupled with the focus on genetic data from individuals of European ancestry, which constrains the applicability of our findings to other diverse populations. Future studies incorporating multi-ancestry cohorts are needed to validate these findings and assess their applicability across different ethnic backgrounds.

In conclusion, our findings demonstrate a role of specific metabolic pathways in the pathogenesis of NDDs, with divergent profiles in AD and PD, which are differently linked with CMD and BMI. These results suggest that genetically adjusted metabolic biomarker profiles can improve early diagnosis of NDDs and guide the development of disease-specific prevention strategies and therapeutic interventions.

## METHODS

### Phenotypic associations

Data from the UK Biobank (application 27412) were utilized for this study. Ethical clearance was obtained from the NHS National Research Ethics Service (reference 11/NW/0382), and all participants provided informed consent. Our main analyses focused on unrelated individuals of White European descent, identified using a KING kinship coefficient threshold of 0.05^69^ who had both Nightingale NMR plasma metabolomics data and complete covariate information available (N = 207,841; mean age = 57.4 years, SD = 8.0; 53.7% female).

Metabolite measures were preprocessed with the ‘ukbnmr’ R package (version 4.4) to mitigate technical artifacts^70^, followed by a rank-based inverse normal transformation^71^. Diagnoses for Alzheimer’s disease (AD) (ICD-10 codes: F00, G30; N=1,746), Parkinson’s disease (PD) (G20–G21; N=1,910), type 2 diabetes (T2D) (E11; N=17,477), coronary artery disease (CAD) (I20–I25; N=25,705), and stroke (I60–I64, G45; N=8,681) were extracted from UK Biobank field 41270. Body Mass Index (BMI) values (mean = 27.4, SD = 4.8) were obtained from field 21001.

For disease phenotypes, logistic regression models were fitted, regressing each outcome on individual metabolites while adjusting for age and sex. Linear regression was applied to BMI with the same covariates.

### Genome-wide association study (GWAS) summary statistics

We incorporated genome-wide association study (GWAS) summary statistics for 249 metabolic biomarkers measured by the Nightingale NMR platform, derived from a meta-analysis performed by our group, which combined data from 207,841 UK Biobank participants and 92,645 individuals from the Estonian Biobank^33^. For neurodegenerative diseases (NDDs) (AD^34,35^, PD^36,37^), cardiometabolic diseases (CMDs) (T2D^38^, CAD^39^, stroke^40^) and BMI^41^, we utilized the latest GWAS summary data restricted to European ancestry cohorts. When possible, we selected datasets excluding UK Biobank samples to minimize overlap and bias in subsequent Mendelian randomization (MR) and conjFDR analyses. All summary statistics were harmonized and quality-controlled using a standardized pipeline (https://github.com/BioPsyk/cleansumstats).

### Linkage disequilibrium score regression (LDSC)

Cross-trait genetic correlations between metabolic markers, NDDs, CMDs, and BMI were estimated using LDSC^72^. Summary statistics were prepared according to LDSC requirements, including variant filtering and removal of SNPs located within the major histocompatibility complex (MHC) region (chromosome 6: 25–35 Mb).

### Bivariate MiXeR

To quantify the extent of shared genetic architecture between each NDDs and each metabolic marker, we applied the bivariate MiXeR framework^44^. MiXeR partitions SNPs into four components: null variants (π_₀_), trait-specific variants (π_₁_ and π_₂_), and shared variants (π_₁₂_). Model selection was guided by the Akaike Information Criterion (AIC), comparing the best-fitting model to two reference scenarios: (i) a baseline infinitesimal model (minimum overlap) and (ii) a model assuming maximum possible overlap constrained by the least polygenic trait. The MHC region was excluded from all GWAS analyses due to the complex LD structure in this region, and chromosome 19 was specifically excluded from the AD analysis due to the strong effect of the *APOE* locus, which can bias estimates of polygenicity^16^.

### Mendelian randomization (MR)

Bidirectional two-sample Mendelian randomization (MR) was conducted using the ‘TwoSampleMR’ R package v.0.6.14.^73^ Genome-wide significant SNPs were identified and pruned for linkage disequilibrium using PLINK (clumping parameters: p=1, r²=0.001, kb=10,000) with the 1000 Genomes European reference panel. Causal effect estimates were obtained primarily via inverse variance weighted (IVW) regression, with weighted median and MR-Egger methods applied to test robustness and detect pleiotropy^74^. Results passing false discovery rate correction (FDR<0.05) in both IVW and weighted median, and nominal significance in MR-Egger, were considered robust.

### Conjunctional False Discovery Rate (ConjFDR)

To identify shared genetic loci jointly associated with NDDs and metabolic markers, we performed conjFDR analyses^45^ using the ‘pleioFDR’ software (https://github.com/precimed/pleiofdr). This approach estimates FDR values conditional on association with the other trait in both directions, taking the maximum as the final conjFDR per SNP. A genome-wide significance threshold of conjFDR<0.05 was applied.

### Gene mapping

Lead SNPs identified by conjFDR were mapped to genes via the Variant-to-Gene (V2G) algorithm implemented by Open Targets Genetic^75^. This approach integrates multiple sources of evidence, including quantitative trait loci (QTL) data, chromatin interaction profiles, computational functional predictions, and proximity to transcription start sites.

### Gene set enrichment

Gene set enrichment analysis was conducted on the genes mapped from conjFDR loci, using the Gene Ontology biological process collection from the Molecular Signatures Database (MsigDB v7.1, c5.bp). For brain tissue, regional gene expression data were obtained from the Allen Human Brain Atlas (https://human.brain-map.org/) based on normalized microarray expression profiles across multiple brain regions from adult donors. For body tissues, we used gene expression data from the Genotype-Tissue Expression (GTEx) Project, version 8 (https://gtexportal.org), which provides RNA-seq– based transcriptomic profiles across 54 human tissues. Enrichment testing utilized Fisher’s exact test via the ‘clusterProfiler’ Bioconductor R package v.4.12.6.^76^. Terms with at least 5 but fewer than 500 mapped genes, and a q-value below 0.05, were considered significant. To reduce redundancy, terms overlapping by more than 80% with more significant sets were filtered out.

### Statistical analyses

All further statistical analyses, data processing, and visualization were performed using R software, v.4.4.1^77^.

## Data Availability

All data utilized in this study were sourced from publicly accessible databases. Access to individual-level data can be requested through the UK Biobank, contingent upon approval from the UK Biobank Access Committee. Summary statistics for the NMR metabolite GWAS have been previously deposited and are publicly accessible via the NHGRI-EBI GWAS Catalog (https://www.ebi.ac.uk/gwas/).

## ACKNOWLEDGEMENTS

This work was partly performed on the TSD (Tjeneste for Sensitive Data) facilities, owned by the University of Oslo and operated by the TSD service group at the University of Oslo, IT Department (USIT). Additional computations were performed on resources provided by UNINETT Sigma2, the national infrastructure for high-performance computing and data storage in Norway.

## AUTHOR CONTRIBUTIONS

S.E.S., D.v.d.M., and O.A.A. conceived the study. S.E.S. performed the Bioinformatics analyses, with conceptual and statistical support from D.v.d.M., A.S., Z.R., and O.A.A. All authors contributed to the interpretation of results. S.E.S. and D.v.d.M. drafted the manuscript, and all authors reviewed and approved the final version.

## SUPPLEMENTARY FIGURES

Suppl Fig 1-5.

## SUPPLEMENTARY TABLES

Suppl Tables 1-11.

