## Supplementary figure 1-5 for "Distinct metabolic signatures of Alzheimer’s and Parkinson’s disease revealed through genetic overlap with metabolic markers"

### SUPPLEMENTARY FIGURES

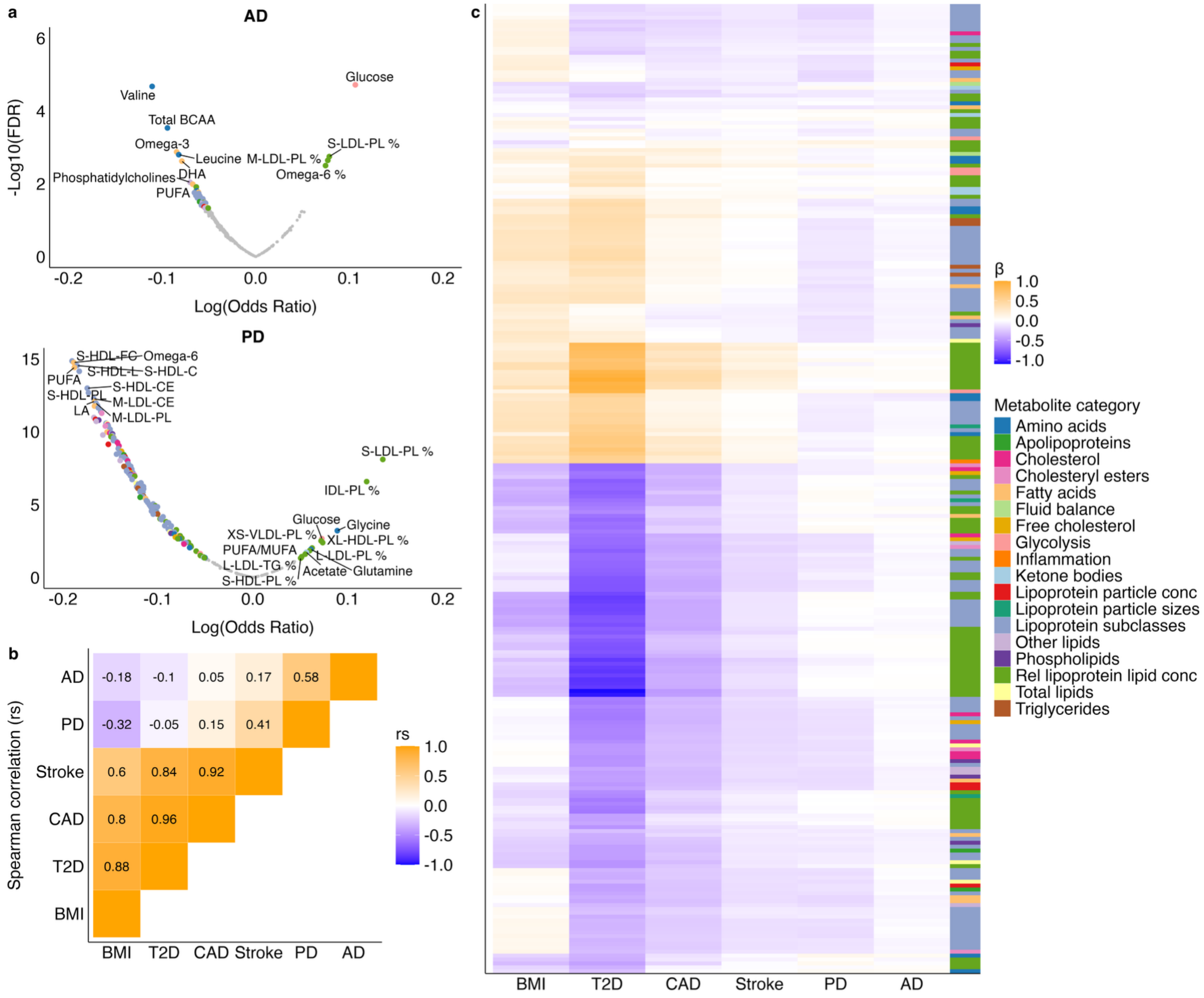


***Suppl Fig 1. Phenotypic associations with metabolic markers, AD, PD and cardiometabolic diseases/traits. a,*** *Volcano plots summarizing the estimated logistic regression coefficients on the x-axis and -log10(p-values) on the y-axis, separately for each NDDs. Significant associations are indicated in color, corresponding to the marker category, and annotated with the marker name, non-significant (FDR > 0.05) in grey.* ***b,*** *A correlation matrix in the top left triangle depicts the Spearman correlation coefficients of logistic regression estimates between these traits across all 249 metabolic markers.* ***c,*** *Heatmap of the regression estimates between the metabolites (y-axis) and cardiometabolic trait/diseases (BMI, T2D, CAD, and stroke) and PD, AD (x-axis), with positive coefficients displayed in orange and negative in blue (see legend). The y-axis is sorted based on hierarchical clustering. The column to the right indicates the metabolic marker categories (see legend). The linear and logistic regression models were adjusted for age and sex. AD=Alzheimer's disease; BMI=body mass index, CAD=coronary artery disease, PD=Parkinson's disease, T2D=type 2 diabetes.*

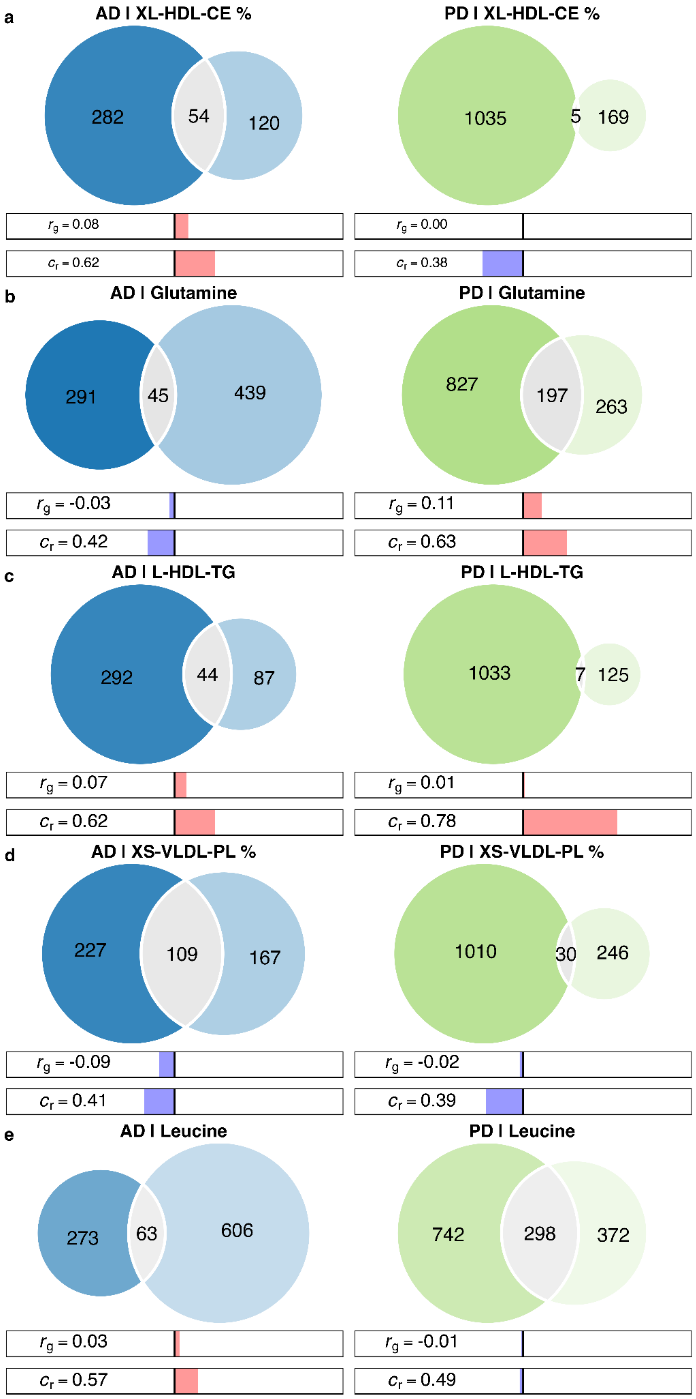


***Suppl Fig 2. Genetic overlap between potential causal metabolic markers for AD and PD, estimated by bivariate MiXeR.*** *Venn diagrams illustrating the genetic overlap between NDDs and **a,** XL-HDL-CE %; **b,** glutamine; **c,** L-HDL-TG; **d,** XS-VLDL-PL %; **e,** leucine. The estimated number of unique and shared variants. Below the Venn diagrams, the bar displays the genetic correlation ($r_g$) and concordance rate ($C_r$; wherein 0.50 indicates an equal number of variants with opposing and same directions of effects on the pair of traits). AD=Alzheimer's disease; NDD=neurodegenerative disease (blue); PD=Parkinson's disease (green).*

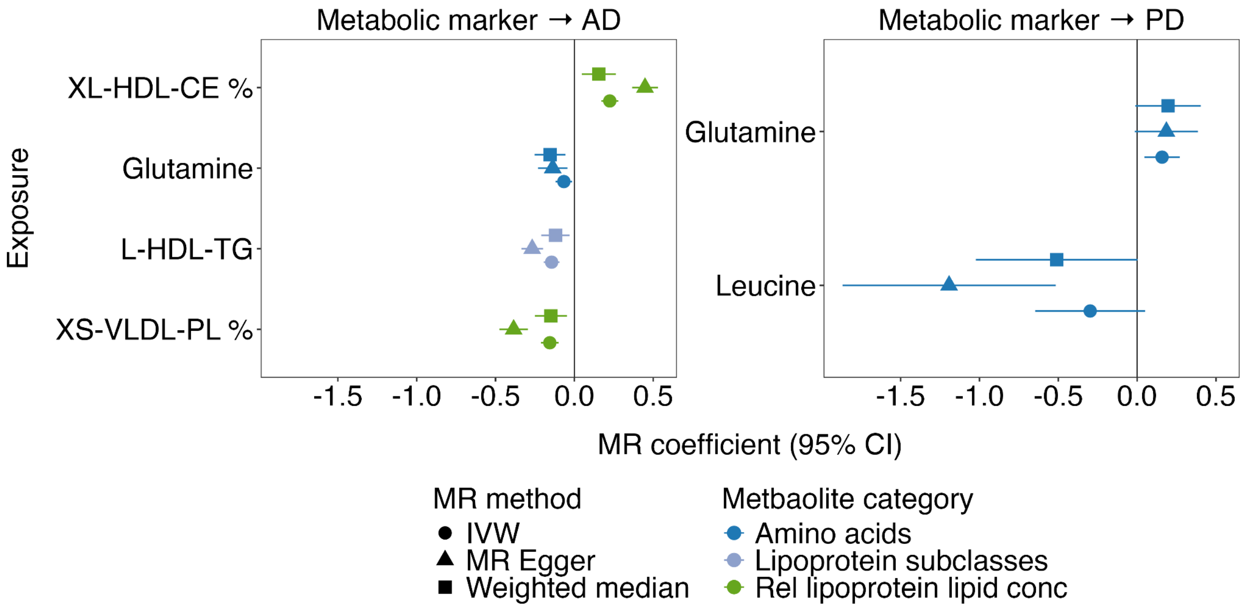


***Suppl Fig 3. Significant causal relationships between metabolic markers on AD and PD, across MR methods. a,*** *The causal effects of the significant metabolic markers on AD and* ***b,*** *suggestively significant for PD, with IVW, MR Egger, and Weighted Median MR coefficients and their 95% confidence intervals (CIs) on the x-axis, the metabolic markers on y-axis. AD=Alzheimer's disease; IVW=Inverse Weighted Median; MR=Mendelian randomization; PD=Parkinson's disease.*

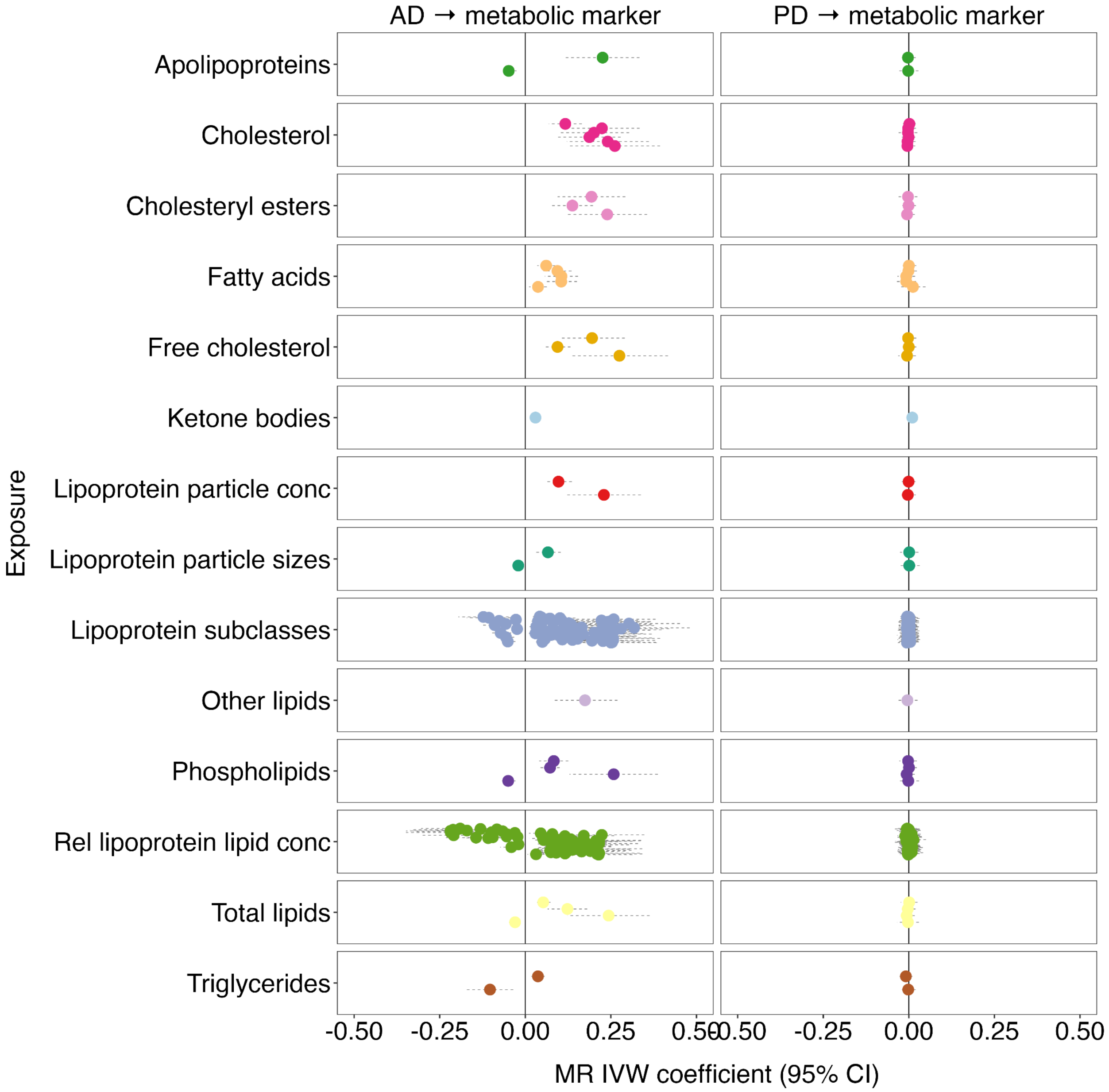


***Suppl Fig 4. Causal relationships of AD and PD on metabolic markers using Mendelian randomization. a,*** *The causal effects of AD and* ***b,*** *PD on metabolic marker categories, with IVW MR coefficients and their 95% confidence intervals (CIs) on the x-axis, the metabolic marker categories on the y-axis. Findings were considered significant if they passed false discovery rate (FDR) correction <0.05 for both IVW and weighted median methods and showed nominal significance in MR-Egger analyses. Only AD showed significant effects on metabolic markers, not PD. AD=Alzheimer's disease; IVW=Inverse Weighted Median; MR=Mendelian randomization; conc=concentration, PD=Parkinson's disease, Rel=relative.*

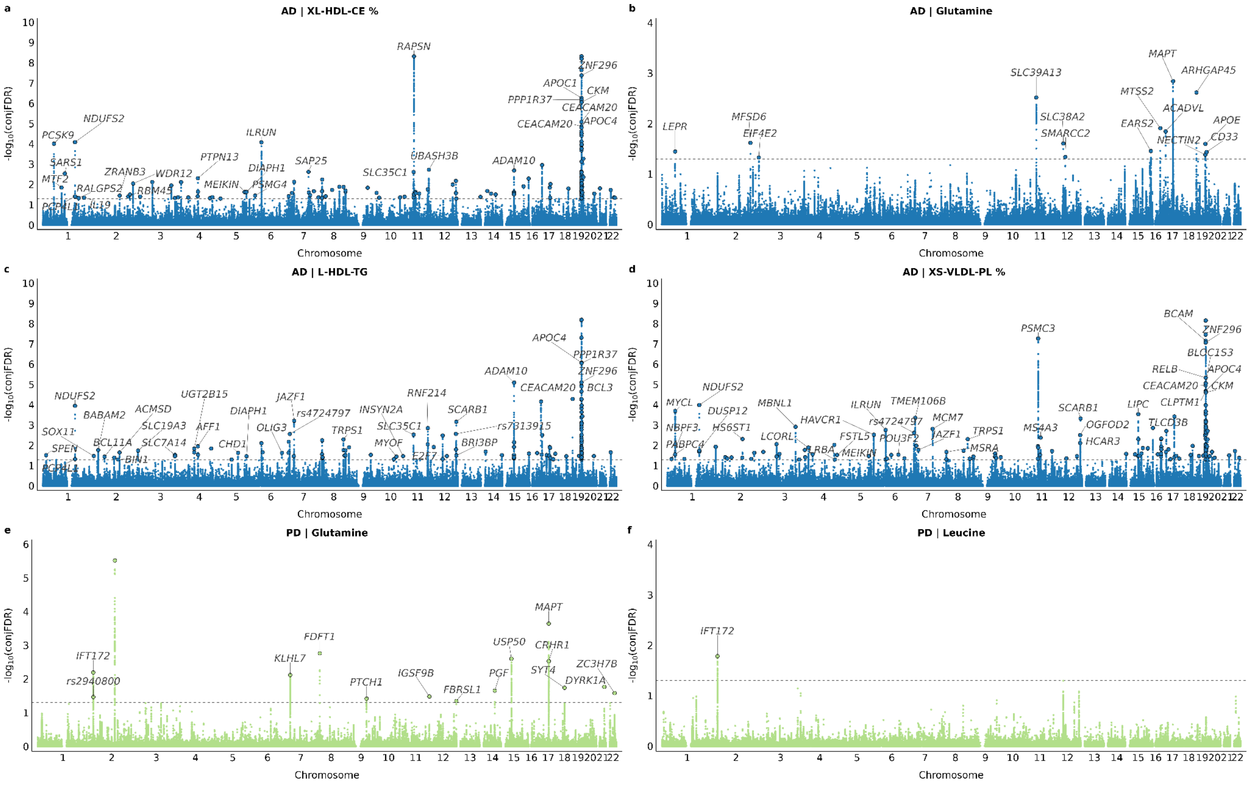

***Suppl Fig 5.*** *Manhattan plots showing the −log10 transformed conjFDR values for each SNP on the y-axis and chromosomal positions along the x-axis for* ***a,*** *AD & XL-HDL-CE %;* ***b,*** *AD & glutamine;* ***c,*** *AD & L-HDL-TG;* ***d,*** *AD & XS-VLDL-PL %;* ***e,*** *PD & glutamine; and* ***f,*** *PD & leucine. SNPs with conjunction FDR<0.05 (i.e., −log10 FDR>1.3) are shown with enlarged data points. A black circle around the enlarged data points indicates the most significant SNP in each LD block. AD=Alzheimer's disease; PD=Parkinson's disease.*
